# Low-dose interleukin-2 in patients with bipolar depression: a phase 2 randomised double-blind placebo-controlled trial

**DOI:** 10.1101/2024.07.19.24310686

**Authors:** Marion Leboyer, Marianne Foiselle, Nicolas Tchitchek, Ryad Tamouza, Roberta Lorenzon, Jean-Romain Richard, Raphaele Arrouasse, Philippe Le Corvoisier, Katia Le Dudal, Eric Vicaut, Pierre Ellul, Michelle Rosenzwajg, David Klatzmann

## Abstract

Immune abnormalities including an insufficiency of regulatory T cells (Treg) and increased blood-based inflammatory markers have been observed in bipolar disorders (BD), particularly during depression. As Tregs are pivotal to control inflammation, Treg stimulation by low-dose IL-2 (IL-2_LD_) could have a therapeutic impact on bipolar depression. We performed a randomized, double-blind, placebo-controlled (2 active: 1 placebo) proof-of-concept trial of add-on IL-2_LD_ in patients with bipolar depression. Patients received a placebo or IL-2_LD_ (1MIU) once a day for 5 days, and then once a week for 4 weeks starting on week 2. The primary objective was to demonstrate a biological Treg response to IL-2_LD_ assessed by fold increase in Treg percentage of CD4+ cells from baseline to day 5. Secondary objectives included safety assessment and mood improvement throughout the study period. This trial is registered with ClinicalTrials.gov, number NCT04133233. Fourteen patients with bipolar depression were included, with 4 receiving placebo and 10 IL-2_LD_. Baseline clinical and biological characteristics were balanced between groups. The primary evaluation criterion was met, with IL-2_LD_ expanding 1.17 [95% CI 1.01-1.34] vs 1.01 [95% CI 0.90 - 1.12] (p=0.0421) and activating Tregs. Secondary evaluation criteria were also met with significant improvements of depressive symptoms and global functioning from day-15 onwards in the IL-2_LD_ treated patients. The treatment was well-tolerated, with no serious adverse events related to treatment. This proof-of-concept trial shows that stimulating Tregs in patients with bipolar depression is safe and associated with clinical improvements. This supports a pathophysiological role of inflammation in BD and warrants pursuing the evaluation of IL-2_LD_ as an adjunct treatment of major mood disorders.

## Introduction

Psychiatric disorders are the largest cause of disability in working-age individuals in Europe, defining a major medical, public health and economic need. In particular, bipolar disorders (BD), characterized by recurrent manic or hypomanic episodes alternating with depressive episodes, are among the top 10 cause of disability. Depressive episodes of BD manifest as decreased energy, loss of pleasure, fatigue, sadness, suicidal thoughts, and cognitive impairments.

BD-caused disability is largely driven by such depressive episodes(Grande et al., 2016) with high suicidal rates and frequent somatic comorbid disorders(Godin et al., 2014). Despite this elevated burden, only a few monotherapies have been approved by regulatory agencies for treating acute depression of BD(Yatham et al., 2018). Antidepressant monotherapy is actually not recommended to treat bipolar depression(Yatham et al., 2018), and even contraindicated due to concerns about inducing manic episodes(Goldberg and Nassir Ghaemi, 2005), reinforcing the need to better understand its specific underlying pathophysiology towards the development of mechanisms-based treatment

There are converging observations linking decreased fitness of regulatory T cells (Tregs) and increased peripheral pro-inflammatory processes - such as high-sensitivity C-reactive protein (hsCRP)/IL-6, brain-derived neurotrophic factor (BDNF)/TNF-α and soluble TNF-α receptor 1 (sTNFR1) - with the pathogeny of BD(Dowlati et al., 2010; Gibney and Drexhage, 2013; Pariante, 2017; Pape et al., 2019). Multiple observations indicate that a Treg insufficiency may contribute to the pathogenesis of BD (do Prado et al., 2013) and of major depressive disorder (MDD) (Ellul et al., 2018) in both mice and humans. In mouse models of depression, Tregs have a protective role against depressive-like behaviour, as exemplified by the association between the decrease of Tregs and onset of depressive-like behaviour(Hong et al., 2013). In humans, adolescents at high risk for mood disorder have a reduced number of Tregs, negatively correlated with their inflammatory state(Snijders et al., 2016). Similarly, several studies have found a decrease in Tregs associated with pro-inflammatory phenotypes(Grosse et al., 2016) in adults with major depressive episodes, including in patients with bipolar depression(Bennabi et al., 2019; Foiselle et al., 2023; Tarantino et al., 2021). Recently, deficits in Tregs have been associated with the severity of depression in patients with MDD(Rachayon et al., 2024). In the same line, epidemiological data, including those from large nation-wide studies(Benros et al., 2013), indicated that patients with BD have a raised prevalence of autoimmune diseases (Vonk et al., 2007).

The concomitant increase in inflammation and decrease in Tregs fitness in MMD is consistent with the fact that one main role of Tregs is to control inflammation. A Treg insufficiency fuelling an inflammation that contributes to MMD pathophysiology suggests that boosting Tregs could be beneficial in MDD. This could be achieved by treatment with low dose IL-2 (IL-2_LD_)(Klatzmann and Abbas, 2015). Indeed, IL-2 is the main cytokine driving Treg development, activation, and proliferation. At low dose, IL-2 is a specific and safe Treg activator, as demonstrated in numerous clinical trials(Saadoun et al., 2011; Koreth et al., 2011; Hartemann et al., 2013; Rosenzwajg et al., 2015a, 2015a; He et al., 2016; Rosenzwajg et al., 2019, 2020; Camu et al., 2020; Humrich et al., 2022; Nigel Leigh et al., 2023; Lorenzon et al., 2024; Barde et al., 2024). These effects are due to the exquisite sensitivity of Tregs to IL-2 that are explained by (i) the constitutive expression of the IL-2 high affinity receptor; (ii) the 20-40 fold higher sensitivity to IL-2 for STAT5 phosphorylation between Tregs and other T cells; (iii) the >100 fold higher sensitivity of Tregs for the gene activation program downstream phosphorylated STAT5 (Rosenzwajg et al., 2015a; Yu et al., 2015),.

In mice, IL-2_LD_ has shown efficacy in the treatment of numerous experimental brain disorders underpinned by neuroinflammation, including in models of multiple sclerosis (MS)(McIntyre et al., 2020), Alzheimer disease (AD)(Alves et al., 2017), ALS(Camu et al., 2020; Nigel Leigh et al., 2023), and brain trauma(Liston et al., 2023). In a murine chronic stress-induced model of depression, IL-2_LD_ corrected the Th17/Treg balance and peripheral tags of low-grade inflammation, and attenuated depression-like behaviours(Liston et al., 2023). These positive effects of boosting Tregs in neuroinflammatory settings could arise from the control of inflammation as well as from an improved tissue regeneration. Indeed, another major role of Tregs is to favour tissue regeneration, as documented in the lung, the muscle and the brain, for example(Mock et al., 2014; Villalta et al., 2014; Becker et al., 2023; Liston et al., 2023).

In humans, there is considerable evidence that Treg stimulation by IL-2_LD_ improves systemic inflammation, including neuroinflammation. Actually, in the first worldwide trial of IL-2_LD_ in an autoimmune disease, we already documented a global anti-inflammatory effect of Treg stimulation (Saadoun et al., 2011). Numerous trials in more than 30 different diseases have demonstrated the excellent safety of IL-2_LD_, including in neuropathological settings such as multiple sclerosis (MS)(Louapre et al., 2023) or amyotrophic lateral sclerosis (ALS)(Camu et al., 2020). Recently, it was further reported that IL-2_LD_ led to significant clinical improvement of patients with ALS (Bensimon et al., 2022; Nigel Leigh et al., 2023), which indicates a positive effect on neuro-inflammation. These data provide a strong rationale for investigating the stimulation of Treg fitness by IL-2_LD_ in bipolar depression. We thus performed a proof of concept, randomized, placebo-controlled trial to test if IL-2_LD_ could stimulate Treg while improving depressive symptoms in bipolar depression.

## Methods

### Study design, participants and treatments

We conceived (ML and DK) the research program on the effects of IL-2_LD_ in depression in 2017 in the context of the H2020 MoodStratification project(Drexhage, 2018). It was then articulated in two parallel trials, one to be conducted in bipolar disorders at the Assistance Publique - Hôpitaux de Paris (DEPIL-2, ClinicalTrials.gov: NCT04133233) and one in both major depressive disorder (MDD) and BD at the Ospedale San Raffaele (IL-2REG; EudraCT N.: 2019-001696-36). IL-2REG was recently reported (Poletti et al., 2024) and here we present the outcomes of the DEPIL-2 trial.

#### Design

DEPIL-2 was a randomized, double-blind, placebo-controlled (2 active / 1 placebo), mono-centre, proof-of-concept trial evaluating the tolerance and the efficacy of IL-2_LD_ in patients with bipolar depression. The primary endpoint was the change in the relative concentration of peripheral blood Tregs at day-5 compared to baseline. Biological secondary endpoints included changes from baseline in Tregs and effector T cells (Teffs) values and activation. Clinical secondary endpoints were from Clinical Global Impression (CGI) and disease-specific scores (MADRS and IDS) at week-6 compared to baseline. Safety was assessed throughout the study (**Figure S1**). DEPIL-2 was registered as “*Low dose IL-2 therapy in patients with a depressive episode in the course of a bipolar disorder*” with EUDRACT number 2018-002777-22 and with ClinicalTrials number NCT04133233.

#### Patients

Patients from the Mondor University hospitals Psychiatry department (Créteil, France) were included if they had (i) a documented diagnosis of a depressive episode according to DSM-5 criteria during a bipolar disorder, (ii) a mild to severe depression (MADRS > 17), (iii a background therapy with a mood stabilizer and/or antidepressant. An additional criterion of having a stable thyroid function was added from patient 12 on. The main exclusion criteria were having severe haematological disorders, vital organ failure, cancer and active HIV, HBV or EBV infections, as well as anti-TPO or anti-TG or anti-TRACKS antibodies from patient 12 on (**Table S1**). The study was approved by the institutional review board of Hôtel-Dieu Hospital in Paris (CPP IDF1) and performed following the Declaration of Helsinki and good clinical practices. Written informed consent was obtained from all participants before enrolment in the study.

#### Treatment

IL-2_LD_ was provided by ILTOO Pharma, Paris, France, as ILT-101®. This is a formulation of adesleukin that is packaged in ready-to-use vials of 1.2 MIU. Treatment administration consists of a first course of daily injections for five days (the induction course), followed by a single injection every week for 4 weeks (the maintenance course; from day15 to day36) (**Figure S1**).

### Randomization and masking

After screening, patients were randomized to the treatment arms (2 active: 1 placebo) using a randomization list generated by computer. All personnel in contact with the patients (research nurses, investigators, clinical research assistants) and assessing outcomes (including laboratory ones) were masked to group assignment until the end of study analyses. Syringes containing placebo and IL-2_LD_ had the same appearance and were labelled according to good manufacturing practice for traceability and accountability purposes. A data safety monitoring board was granted access to unmasked data but had no contact with the study participants. The unmasked pharmacists had no contact with study participants.

### Patients monitoring

Patients were clinically monitored in the Mondor University hospital’s Clinical Investigation Center (CIC) by a trained psychologist at each visit (screening, inclusion, day 0-5, 15, 22, 29, 36, 60). Depressive symptomatology was evaluated by the Montgomery-Åsberg Depression Rating Scale (MADRS) and the Inventory for Depressive Symptomatology (IDS-C)(Baer and Blais, 2010). Manic symptomatology was assessed using the Young Mania Rating Scale (YMRS)(Baer and Blais, 2010). Anxiety was evaluated by the Hamilton rating scale for Anxiety (HAM-A)(Baer and Blais, 2010), current suicide risk with the Columbia – Suicide Severity Rating Scale (C-SSRS)(Baer and Blais, 2010), and global functioning with the Clinical Global Impression (CGI)(Busner and Targum, 2007). Psychotic symptoms (PANSS)(Baer and Blais, 2010) and self-reported anhedonia (SHAPS)(Baer and Blais, 2010) were assessed at day 0 and day 60. All scales were French-validated versions. All patients remained under the same mood stabilizers and/or antidepressants before and all along the trial; this psychiatric treatment had been decided by the patient’s psychiatrist. Any unexpected change in psychiatric medication was recorded by a trained physician.

### Immunomonitoring

Blood samples were obtained for immunological tests at screening, day 1 (baseline), day 5 and day 60. All blood samples were acquired immediately before the administration of IL-2_LD_. All the immunomonitoring procedures (flow cytometry and quantification/analysis of cytokine and chemokine expression levels) were performed as previously described(Louapre et al., 2023; Rosenzwajg et al., 2019) (Supplementary methods).

### Outcomes

The primary objective was to demonstrate the Treg response to IL-2_LD_ in patients with bipolar disorders experiencing a depressive relapse. The primary endpoint was the Treg (% of CD4+ T cells) fold increase compared to baseline at D5. The secondary objectives were to assess (i) the safety of IL-2_LD_ in this population by describing the frequency and type of adverse events all along the study, (ii) the improvement of immune homeostasis (including Treg activation) and (iii) symptomatic assessment of mood improvements using MADRS, IDS-C, YMRS, HAM-A, C-SSRS, and CGI.

### Statistical analysis

The global between-group differences in the time-dependent profile of changes in Tregs and clinical scores or immunological parameters were analysed throughout the treatment period by non-parametric ANCOVA for repeated measurements, using baseline values as covariable (Conover method). According to their statistical distribution, other quantitative criteria were compared using a Student t-test or Mann-Whitney test. All the tests were performed using SAS version 9.4 (from the SAS Institute).

The Wilcoxon test was employed to identify which clinical scales and subscales were altered over time in patients treated with IL-2 and those receiving a placebo. To measure the extent of these changes, the Cliff’s Delta effect size was utilized, which varies from -1 to 1. A value of 0 signifies no significant change, while -1 and 1 indicate systematic diminution and augmentation, respectively, of the clinical scales and subscales scores. Cliff’s Delta was used in the volcano plot to quantify the magnitude of change because it effectively measures the variation in clinical scales that range with integral values, including zero. This method is particularly relevant as it avoids the pitfalls of fold-change representation, which can produce misleading values when dealing with zero or near-zero data points. By using Cliff’s Delta, we provide a more accurate and interpretable measure of effect size for the observed changes in clinical outcomes.

## Results

### Study design and patient characteristics

Between June 2020 and May 2022, 20 patients with ongoing bipolar depressive episode disorder were assessed for eligibility, of which 17 were included and 14 randomized (**Figure 1**). The study design provided for two patients to be treated with IL2_LD_ for one patient receiving a placebo. Altogether, four patients received the placebo and 10 received IL-2_LD_. One patient dropped out on day-15 for a biological hypothyroidism without clinical symptoms.

**Figure 1.**
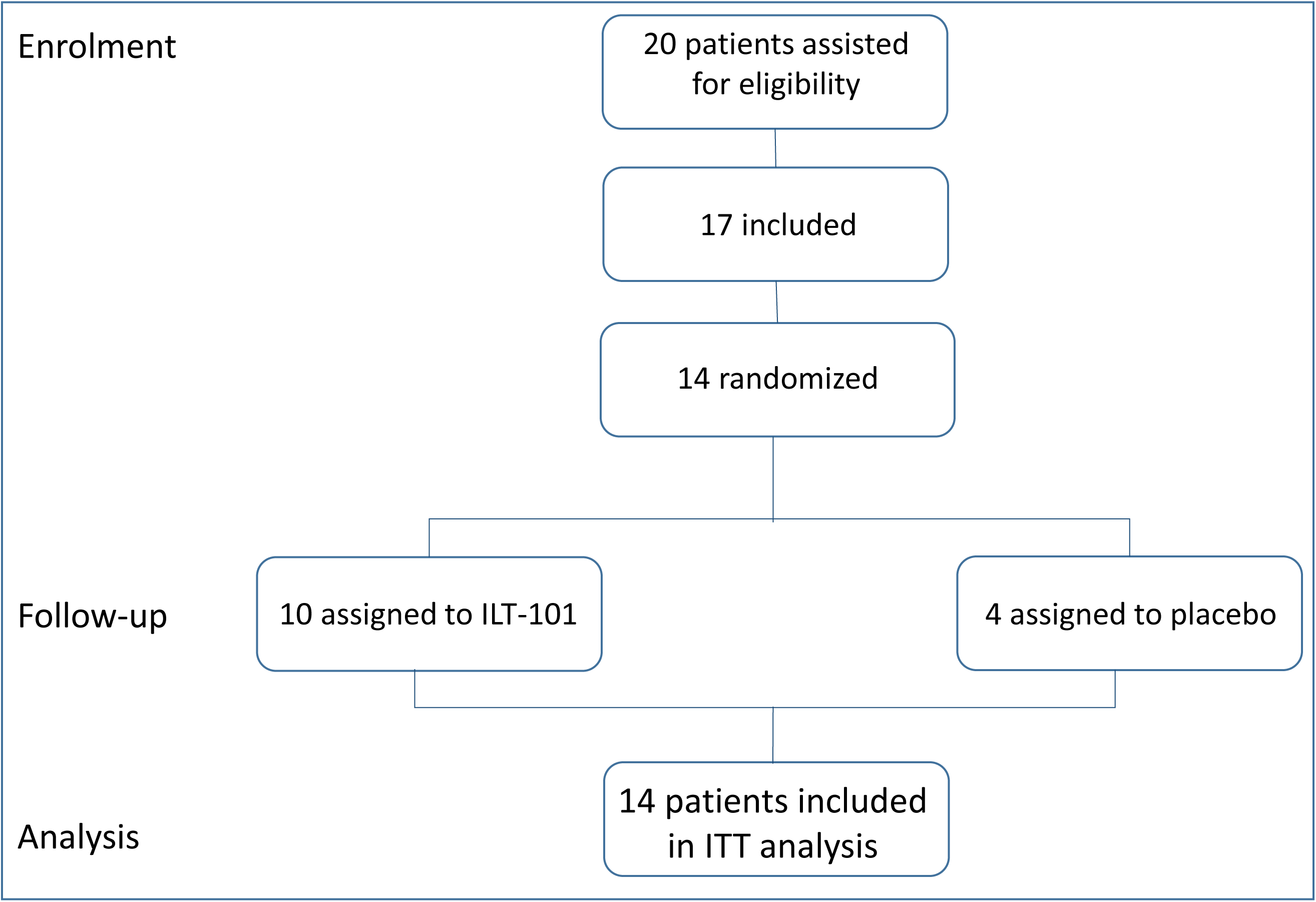
Trial Profile.

Patients’ characteristics are reported in **Table 1**. There were 14 females and 1 male, whose mean age was 48.07 years (±6.96). As assessed by the different specific scales, patients had moderate depressive symptoms at baseline (MADRS = 26.64 ± 5.89, IDS-C = 35.214 ± 9.633, HAM-A = 17.786 ± 6.375, PANSS = 45.071 ± 6.342 and a CGI = 5.21 ± 0.89) without any manic symptom (YMRS = 1.429 ± 1.399). They had a normal-to-moderate level of anhedonia (SHAPS = 4.929 ± 3.668). Their mean Tregs percentages at baseline was 8.293 (±1.717). Patients’ clinical and biological characteristics did not show any imbalance between the placebo and IL-2 groups (**Table 1**).

**Table 1.** Demographic and baseline clinical, laboratory characteristics of randomized patients.

| Patient characteristics | ILT-101<br>(N=10) | Placebo<br>(N=4) | Total<br>(N=14) |
| --- | --- | --- | --- |

| **Demographic characteristics** |  |  |  |
| --- | --- | --- | --- |
| Male (n, %) | 0 (0%) | 1 (25%) | 1 (7%) |
| Female (n, %) | 10 (100%) | 3 (75%) | 13 (93%) |
| Age (mean±SD) | 50.6 ± 5.370 | 41.5 ± 6.801 | 48.00 ± 7.121 |
| **Clinical characteristics**<br>(Mean±SD) | **ILT-101**<br>(N=10) | **Placebo**<br>(N=4) | **Total**<br>(N=14) |
| MADRS | 26.200 ± 4.367 | 27.750 ± 9.535 | 26.643 ± 5.891 |
| YMRS | 1.400 ± 1.265 | 1.500 ± 1.915 | 1.429 ± 1.399 |
| IDS-C | 33.300 ± 7.088 | 40.000 ± 14.445 | 35.214 ± 9.633 |
| HAM-A | 15.700 ± 3.683 | 23.000 ± 9.201 | 17.786 ± 6.375 |
| PANSS | 44.100 ± 5.425 | 47.500 ± 8.660 | 45.071 ± 6.342 |
| SHAPS | 4.500 ± 4.035 | 6.000 ± 2.708 | 4.929 ± 3.668 |
| CTQ | 41.300 ± 11.304 | 58.500 ± 16.299 | 46.214 ± 14.656 |
| CGI | 5.100±0.738 | 5.500±1.291 | 5.214±0.899 |
| **Laboratory characteristics** |  |  |  |
| % Tregs at Baseline (mean± SD) | 8.661±1.831 | 7.373±1.066 | 8.293±1.717 |

Abbreviations: MADRS: Montgomery-Åsberg Depression Rating Scale; YMRS: Young Mania Rating Scale ; IDS: Inventory for Depressive Symptomatology ; HAM: Hamilton Anxiety Rating Scale ; PANSS: Positive and negative syndrome scale ; SHAPS: Snaith-Hamilton pleasure scale ; CTQ: Childhood Trauma Questionnaire ; CGI: clinical global impression

### Biological response to IL-2_LD_

At baseline, the mean ± SD percentage of Tregs in CD4^+^ T cells was 8.66±1.83 in the IL-2_LD_ treated group and 7.37±1.07 in placebo-treated group (p-value=ns) (**Table 1**). At day-5, there was a significant increase in the Treg percentage fold change in the IL-2_LD_ vs placebo groups (Primary criteria: 1.17±0.23 vs 1.0±0.07, p-value=0.042) (**Figure 2A**). At day-60, a significant Treg percentage increase was maintained in the treated group (**Figure 2A**). As there were no effects of IL-2_LD_ on effector T cells, this translated into a significantly increased Treg/Teff ratio at day-5 and -60 (**Figure 2B**).

**Figure 2.**
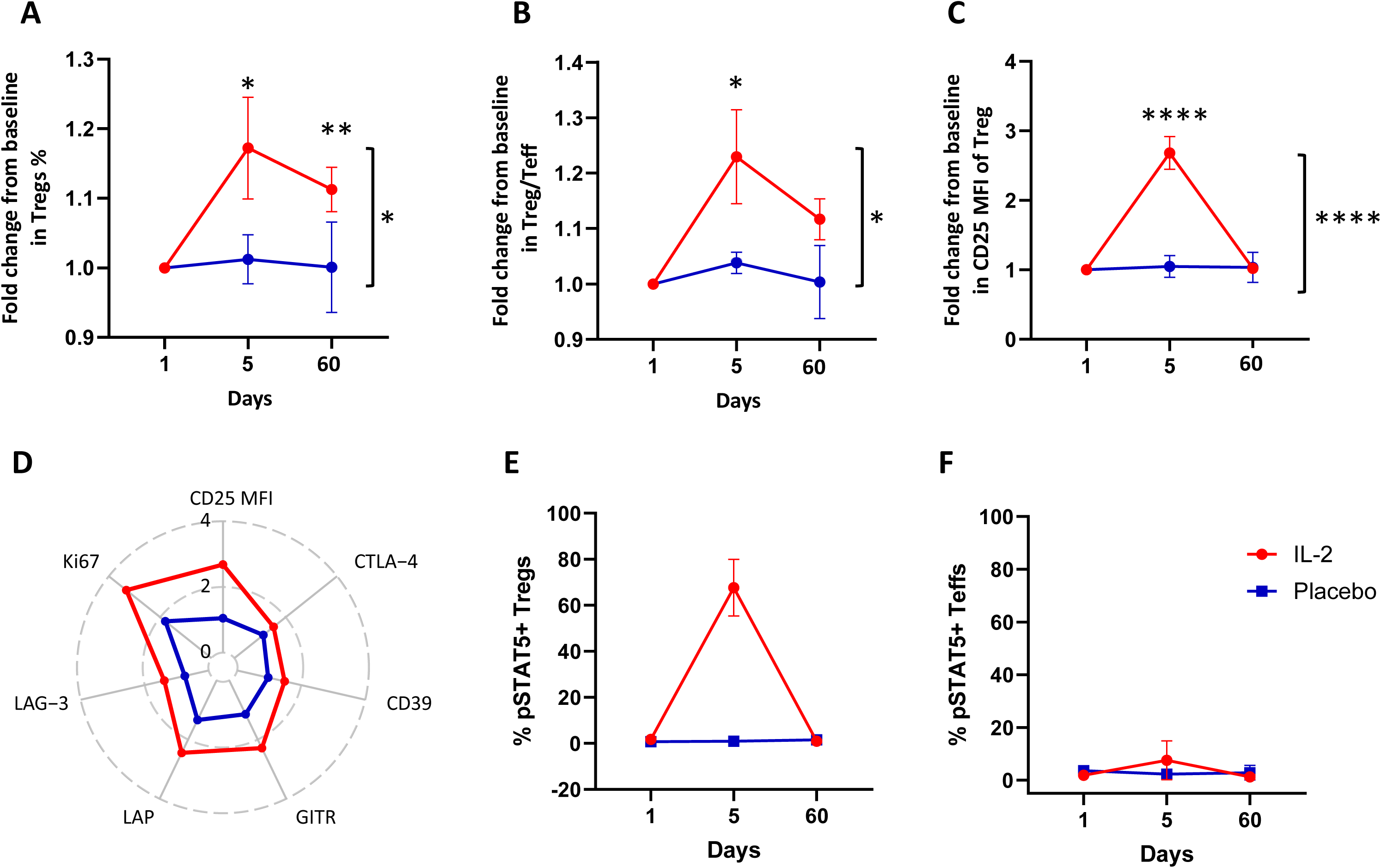
Quantitative and qualitative changes in Tregs and Teffs. Treg cells were gated in CD4^+^ T cells and identified as CD25^hi^CD127^lo/-^Foxp3^+^cells. **(A-F)** Changes represented as mean ± sem by patient groups (IL-2 in red and Placebo in blue) from day 1 to day 60, in **(A)** Tregs as percentages among CD4+ T cells; (B) Treg/Teff ratio; **(C)** CD25 MFI in Tregs; **(D)** Functional/activation markers expression by Treg; **(E, F)** Basal STAT5 phosphorylation of treated and placebo patients’ Treg and Teff. The global differences between the time-dependent profile of changes between treated and placebo patients throughout the treatment period was analysed by non-parametric ANCOVA for repeated measurements, using baseline values as covariable (Conover method). Comparisons of each parameter in treated patients at day 5 and day 60 were made using t-test or the Mann-Whitney test according to their normality distribution. (*p<0.05; **p<0.01; ***p<0.001).

IL-2_LD_ also activated the expanded Tregs. At day 5, there was a highly significant increase of CD25 (**Figure 2C**), a classical marker of IL-2-induced Treg activation(Rosenzwajg et al., 2015b, 2019, 2020; Louapre et al., 2023). Of note, the day-five sampling occurs 24 hours after the previous IL-2 injection, a time at which the activation markers are still increased. The day-60 sampling occurs 24 days after the last IL-2_LD_ injection and, as expected, the activation markers have returned to baseline. There was also a similar increase of classic Treg activation markers (CTLA4, GITR, CD39, and LAP) and of Ki67 (**Figure 2D**). In line with the increase of activation markers, the basal levels of phosphorylated stat5 (pStat5) were also elevated, indicating a robust activation of Tregs (**Figure 2E**). There was no such activation of Teffs (**Figure 2F**). Moreover, as previously described (Rosenzwajg et al., 2019, 2015b), we observed an increased frequency of the regulatory CD56^bright^ NK cell subset whiles we did not observed modification in major lymphocytes subsets or in eosinophils(**Figure S2A and S3**). All these markers returned to baseline value at the end of the follow-up (**Figure 2 E-F and Figure S2A and S2B**).

### Clinical response to IL-2_LD_

We performed a global analysis of all the clinical scales and subscales monitored over time in both IL-2-treated and placebo-treated patients (**Figure 3A and Figure S4**). We represented these scales and subscales changes in terms of both magnitude (x-axis) and statistical significance (y-axis) using volcano plots. The clinical scales and subscales that were significantly modified (i.e. p-value<0.01) relative to baseline are coloured in green and labelled (**Figure 3A**). Noteworthy, there were no modifications of any of the parameters in the placebo-treated patients. In marked contrast, there was a significant decrease in clinical scales or subscales in the IL-2-treated patients. These improvement started on day-15, and remarkably their number, intensity and significance increased over time. The improved scales and symptoms encompassed the Clinical Global Impressions (CGI) scale, the Hamilton Anxiety Rating Scale (HAM-A), particularly depressed mood (HAM_A_06), and various aspects of the Inventory for Depressive Symptomatology (IDS), including expressions of sadness (IDS_05), mood reactivity (IDS_08), mood quality (IDS_10), pessimistic outlooks on the future (IDS_17), and aspects related to physical energy or leaden paralysis (IDS_30). Additionally, the Montgomery-Åsberg Depression Rating Scale (MADRS) indicated notable changes, specifically in the areas of apparent sadness (MADRS_01), reported sadness (MADRS_02), and lassitude (MADRS_07). A Principal Component Analysis based on all these parameters at baseline, day-36 and day-60 showed that the baseline values were well separated from most of the day-36 and day-60 values in the IL-2 treated patients **Figure S5A**) and that CGI, HAM, IDS, and MADRS contributed most to this separation (**Figure S5B)**.

**Figure 3.**
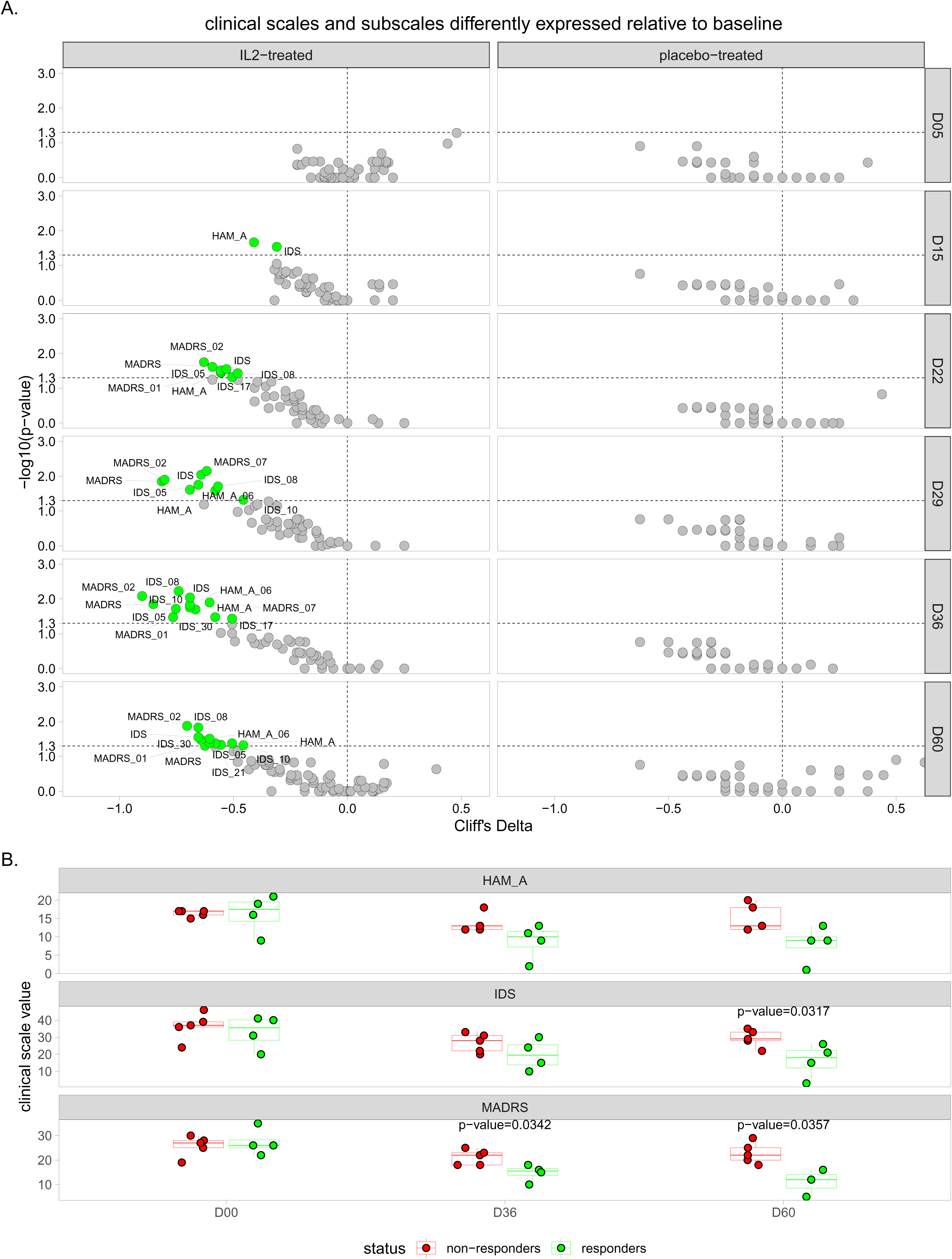
Global analyses of clinical changes along the treatment period. (**A**) Volcano plot representations of clinical scales and subscales modified over time in the IL-2 _LD_-treated and placebo-treated patients. The x-axis in the representations corresponds to the magnitude of the changes, shown as Cliff’s delta values. The y-axis in the representations corresponds to the statistical significance of changes, shown as -log10 of the p-value. Clinical scales and subscales significantly modified relative to baseline are colored in green and labelled. **(B)** Jitter and boxplot representations showing the HAM-A, IDS, and MADRS scales changes between responders and non-responders at baseline, D36, and D60 post-beginning of the IL-2_LD_ treatment. The p-values are indicated for statistically significant comparisons.

Noteworthy, the HAM, IDS, and MADRS scores tend to improve along time in the IL-2 but not in the placebo groups (**Figure S5C).** In a post-hoc exploratory analysis defining responder patients in the IL2 group as those with a decrease of 2 or more points in the Clinical Global Impressions (CGI) scale at D60 relative to baseline, we observed significant decreases of the IDS and MADRS scores, and a trend for decrease of the HAM score, at day 36 and Day-60. (**Figure 3B).**

### Safety

Safety was assessed as immediate tolerance after injection and adverse events occurring between visits. There was one reported severe adverse event (AE) affecting on patient in the IL2 arm presenting tremors not being assigned as related to treatment. Non-severe AEs were reported at least once in 50% of our patients, all in the IL-2_LD_ group. One patient developed a biological hypothyroidism without clinical symptoms and dropped out of the study on day 15. Other adverse reactions consisted of injection site reactions, which resolved within 24 hours (**Table S2**).

## Discussion

We performed a proof of concept, randomized, placebo-controlled trial to test if IL-2_LD_ could stimulate Treg while improving depressive symptoms in bipolar depression. The primary evaluation criterion was met, with IL-2_LD_ expanding and activating Tregs. Secondary evaluation criteria were also met with significant improvements of depressive symptoms and global functioning from day-15 onwards in the IL-2_LD_ treated patients. The treatment was well-tolerated, with no serious adverse events related to treatment.

As there were no previous indications that IL-2_LD_ indeed could stimulate Tregs in this population, the primary criterion for the trial was the capacity of IL-2_LD_ to expand Treg. The primary outcome was met, with a modest 20% increase in Treg percentage at day-5 that however underestimates the IL-2_LD_ effects. Indeed, Indeed, Day-5 does not capture the peak of the Treg response to the induction course, which is around day-8(Hartemann et al., 2013 and unpublished observation). The IL-2_LD_ effects on Tregs are further evidenced by a coordinated increase of the main Tregs activation markers, including the unstimulated basal levels of phosphorylated Stat.5 that is the best surrogate marker of an IL-2/IL-2receptor interaction. Thus, IL-2_LD_ properly expands and activate Tregs in depressed BD patients, allowing to assess its potential effects of clinical depression.

There were no significant changes in any of the clinical parameters of depression in the placebo-treated group. In contrast, despite the small size of the IL-2_LD_ treated population, a gradual improvement of the depressive symptoms was observed in the IL-2_LD_ treated group, concomitant to the improved Treg fitness in this population(Klatzmann and Abbas, 2015). These improvements were numerous and coherent. First, as expected, no effects were seen at day-5. Then, starting on day-15, the number, intensity, and significance of the changed parameters increased over time. In detail, we observe an early response focused on improved mood and sadness, which is consistent across scales and strengthens over time. From D30 onwards, there was an improvement in physical parameters such as energy. . The co-analysis of these parameters by PCA could remarkably separate the pre- and post-treatment time points. CGI, IDS, IDS and MADRS scales and subscales are the most important contributors to this separation. Importantly, the robustness of this separation is asserted by having >75 % of the data variance being represented on the first axis. In contrast to study in other diseases(Humrich et al., 2022; Rosenzwajg et al., 2020), there was no correlation between Treg numbers, percentages, or activation markers and the clinical response, which we attribute to the known variability in Treg measurements(Hartemann et al., 2013) and the small size of the study. Despite, we are confident that the antidepressant add-on effect of IL-2_LD_ is related to the Treg activation, given the concomitant increase of all the makers of Treg activation and efficiency(Louapre et al., 2023; Rosenzwajg et al., 2015b, 2019, 2020; Humrich et al., 2022; Lorenzon et al., 2024).

In line with these positive effects of IL-2_LD_ on the depression of BD patients, we also observed improvements of depression in patients with various types of autoimmune diseases that we treated with IL-2_LD_. Specifically, we observed marked improvements in the evolution of symptoms of depression in patients with SLE treated with IL-2_LD_ (Ellul et al, in preparation).

Altogether, our results support that the known anti-inflammatory effects of Treg stimulation(Saadoun et al., 2011) contribute to improve depression. However, besides its effects on inflammation, Tregs stimulation by IL-2 could also have direct effects on the brain. Indeed, IL-2 stimulated Tregs promote survival and neurite extension of cultured cortical, hippocampal, septal, striatal, and cerebellar neurons(Hanisch UK, 1995). These effects are thought to be mediated by the inactivation of the Glycogen synthetase kinase-3-beta (GSK-3β) axis(Braunstein et al., 2008) which constitute one of the main targets of Lithium, the gold standard mood stabilizers for the treatment of BD, and a preferential target for new drug development in the treatment of bipolar depression(An et al., 2012). Increased GSK3-3β activity has been reported in post-mortem brain tissues of depressed suicide victims and in peripheral tissues of patients with bipolar depression.

Blocking GSK-3β should thus promote neuroplasticity and oligodendrocyte homeostasis. Actually, an interplay between inhibition of GSK-3β and increased IL2(Ohteki et al., 2000) has already been reported and inhibition of GSK-3β has been shown to induce the generation and suppressive function of Tregs(Cheng et al., 2020). Thus, IL-2_LD_ might act synergistically with both mood stabilizers and antidepressants to mediate a GSK-3β-mediated rescue the impaired neuroplasticity associated with depression(Machado-Vieira et al., 2014) and promote the repair of Grey Matter and White Matter damages associated with mood disorders, in particular in bipolar depression.

The small sample size of our study presents limitations in terms of statistical power and generalizability of the findings. As a proof-of-concept trial, our primary goal was to establish preliminary evidence for the efficacy and safety of IL-2_LD_ in treating bipolar depression. However, the limited number of participants may reduce the robustness of our results and increase the risk of type II errors. However, our results are in line with the recently published data of the IL-2REG trial that was conducted in parallel within the same European project (Poletti & al). This study that was designed together with ours used the same treatment scheme for patients with Major Depressive Disorder and patients with bipolar depression, while we have only treated patients with bipolar depression in ours. Both studies report a rapid increase in Tregs, followed by a later induction of an antidepressant effect that persisted in the month after the end of the trial. Collectively, this highlights the robustness of our observations.

In conclusion, our POC trial reports a striking concomitant Treg stimulation and clinical response in patients with bipolar depression that reinforces the notion that inflammation has a direct role in bipolar depression (Drexhage, 2018; Poletti et al., 2024). As the treatment is safe, as already reported in more than 25 other diseases(Raeber et al., 2023), our results warrant to pursue properly powered trial to demonstrate the therapeutic efficacy of IL-2_LD_ in bipolar depression, and beyond in mood disorders in general.

## Data Availability

All data produced in the present study are available upon reasonable request to the authors

## Authors’ contribution to the current study

ML and DK conceived and supervised the study. EV, MR, ML and DK designed the study.

MF performed the clinical follow-up of patients under the supervision of ML and RT. JRR contributes to the data management of the study.

PLC, KLD assumed patients’ management during the study. CB oversaw the biobank.

MR supervised the immunomonitoring.

NT, RT, MR and DK analysed the biological results. NT, ML, MF, PE and RL analysed the clinical results.

ML, RT, NT, RL, PE, MR and DK wrote the first draft of the manuscript that was edited by all authors.

## Acknowledgement

The research program on the effects of IL-2_LD_ in depression was defined by ML and DK in 2017 as part of the H2020 MoodStratification proposal (Coordinator: H.Drexhage, 2018), and was later joined by Francesco Benedetti from Ospedale San Raffaele, Milan, Italy. We thank Hemo Drexhage and Francesco Benedetti for stimulating discussions. DEPIL-2 was primarily funded by MOODSTRATIFICATION. We thank ILTOO pharma for provided the study drugs. The immunomonitoring was supported by the ANR grant iMAP (ANR-16-RHUS-0001) to DK and ANR grant I-GIVE (ANR-13-SAMA-0004-01) to ML. We are grateful to the patients for their participation. We thank the personnel of the Pitié-Salpêtrière Clinical Investigation Center for Biotherapies (CIC-BTi), Michèle Barbié, Natalie Féry, Catherine Ferrapie, Nadia Graffin, Aurélie Marc, Alexandra Roux and Fabien Pitoiset for their excellent assistance and Wahid Boukouaci, Ching-Lien Wu, Jihène Bouassida from the Translational Neuropsychiatry laboratory and Caroline Barau from the Plateforme de Ressources Biologiques for their excellent technical contribution.

