## Supplemental materials for "Low-dose interleukin-2 in patients with bipolar depression: a phase 2 randomised double-blind placebo-controlled trial"

### Appendix

### TABLE OF CONTENT


- Table S1: Inclusion/Exclusion criteria
- Table S2: Summary of adverse events
- Figure S1: Study design
- Figure S2: Changes in CD56bright NK cells upon ld-IL2 treatment and in Tregs' activation markers at day 60
- Figure S3: Changed in Eosinophils and major lymphocytes populations
- Figure S4 – Heatmap representation of clinical scales and subscales modified along the time course of the study.
- Figure S5: Multivariate analysis of clinical changes during the treatment period and comparison with placebo-treated individuals
- Supplementary Methods


**Table S1: Inclusion/Exclusion Criteria**

| Inclusion Criteria | Exclusion Criteria |
| --- | --- |
| - A depressive episode according to DSM-V criteria in the course of a bipolar disorder<br>- MADRS > 17<br>- Age 18-65 years;<br>- Already on a mood stabilizer and/or antidepressant<br>- patient with a normal or controlled thyroid function<br>- Male or female both using effective methods of contraception during treatment if sexually active.<br>- Specifically; Females (if sexually active) with childbearing potential must use contraceptive methods that are considered as highly-effective (pearl index < 1). The following methods are acceptable: Oral, injectable, or implanted hormonal contraceptives (with the exception of oral minipills ie low-dose gestagens which are not acceptable (lynestrenol and norestisteron), Intrauterine device, Intrauterine system (for example, progestin-releasing toit),<br>- Signed informed consent, able to understand, speak and write the national language. | - Contraindication to IL-101 therapy:<br>- Hypersensitivity to active substance or excipient;<br>- Active infection requiring antibiotics therapy;<br>- Organ failure (e.g., liver, kidney, lung and heart);<br>- Immunosuppressed patient<br>- Hepatotoxic, nephrotoxic, myelotoxic or cardiotoxic drugs<br>- Other chronic diseases<br>- Signs of active infection requiring treatment<br>- Previous history of organ transplantation<br>- Leukocytes < 4000 / mm3, platelets < 100 000 / mm3, Hemoglobin < 10.0 g/dL or 6.2 mmol/L, red cell blood < 3.5 T/L.<br>- Anti-TPO or anti-TG or anti-TRACKS positive at inclusion.<br>- Use of anti-inflammatory medication on a regular basis for a chronic inflammatory/autoimmune Disorder (NSAD, immunosuppressant IV-Ig based treatment);<br>- Ongoing fever >38°C<br>- uncontrolled type I or II diabetes;<br>- Existing cancer or history of cancer in the last 5 years (except skin epidermoid cancer or in-situ cervix cancer);<br>- Existing or planned pregnancy or lactation;<br>- Person under legal protection (1121-8 of CSP, Public Health Code<br>- Pregnant and parturient and Breast-feeding women (1121-5 of CSP)<br>- legally detained person (1121-6 of CSP)<br>- hospitalisation without consent |

| | |
|---|---|
| | - under the age of majority (1121-7of CSP)<br>- Immediate risk for suicidal behaviour (MADRS-item 10 >2 or columbia > 2 for suicide idea);<br>- Known HIV infection or clinically manifest Acquired Immune Deficiency Syndrome (AIDS), Parkinson's or Alzheimer's disease, or any other serious condition likely to interfere with the conduct of the trial;<br>- Participation to an interventional study concomitantly or within 30 days prior to this study, except in the cohort studies aiming at the analysis of immuno-inflammatory biomarkers and/or brain imaging studies.<br>- Patients thought to be unreliable or incapable of complying with the requirements of the protocol;<br>- Patient is relative of, or staff directly reporting to the investigator;<br>- Patient is employee of the sponsor. |

**Supplementary table 2. Summary of adverse events**

| | **ILT-101**<br>N° Event<br>(N° Patients)<br>[% administrations] | **Placebo**<br>N°Event<br>(N° Patients)<br>[% administrations] | **Overall population**<br>N° Event<br>(N° Patients)<br>[% administrations] |
|---|---|---|---|
| **Treatments administered** | 86 (10) | 45 (4) | 131 (14) |
| **Serious Adverse events (SAE)** | 1 (1) | 0 | 1 (1) |
| **Non Serious Adverse events (NSAE)** | 40 (7) | 0 | 40 (7) |
| **NSAE related to treatment** | 39 [45,3%] | | 39 [29,7%] |
| **Most common NSAE related to treatment** | | | |
| Injection-site reaction | 38 [44,2%] | | 38 [29,0%] |
| Others NSAE<br>*Asthenia*<br>*Persistent severe tremors* | 2 [2,3%]<br>*1 [1,2%]*<br>*1 [1,2%]* | | 2 [1,5%]<br>*1 [0,8%]*<br>*1 [0,8%]* |

#### Supplemental Figures

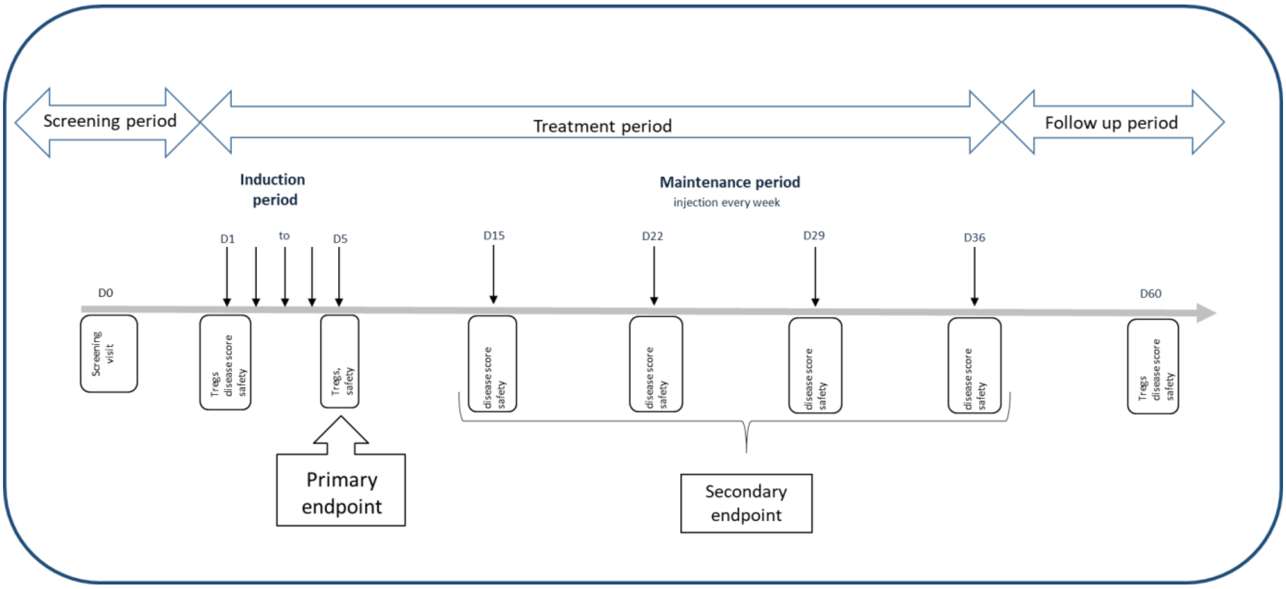


**Figure S1: Study design**

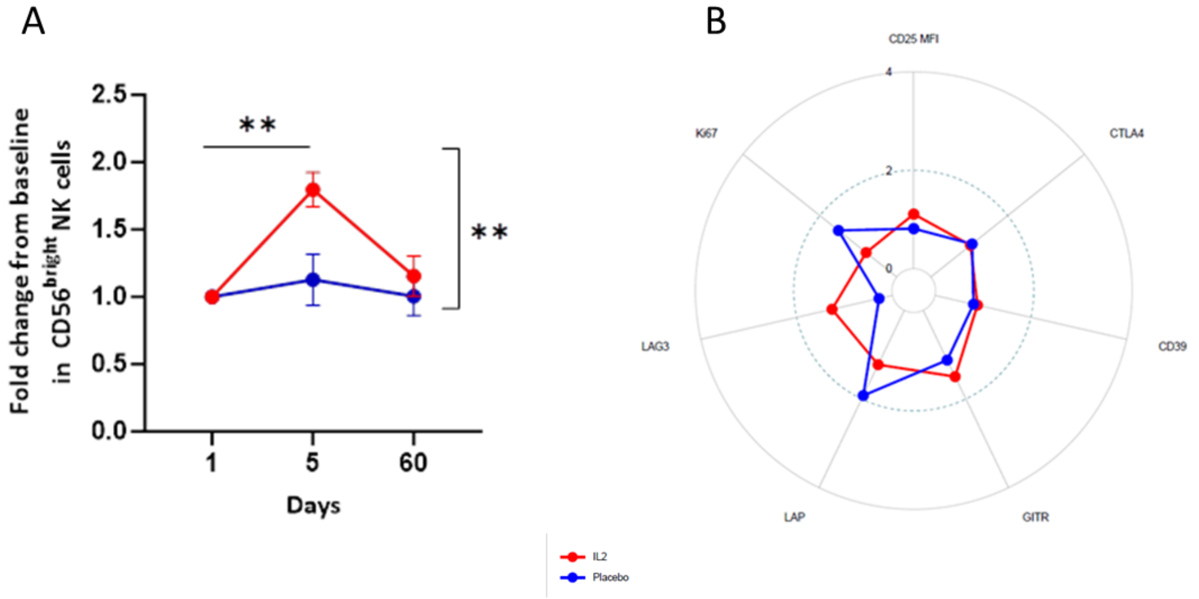


**Figure S2**: A) Changes in CD56$^{bright}$ NK cells upon ld-IL2 treatment; B) Functional/activation markers expression by Treg at day 60 shown as fold change from baseline

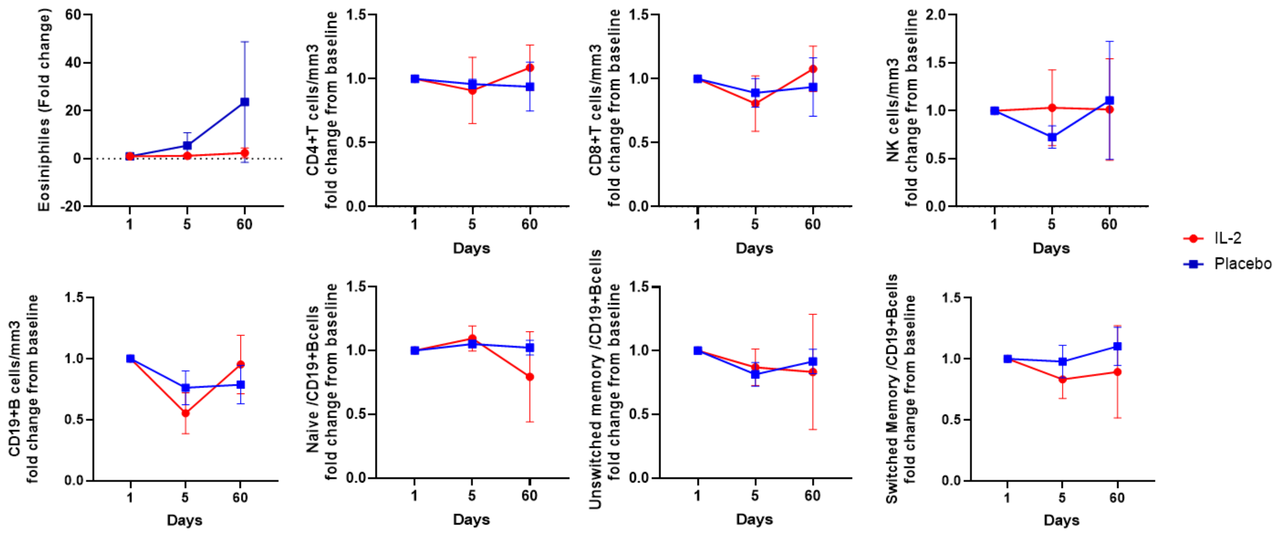


**Figure S3**: Changes in Eosinophils, CD4$^{+}$ T cells, CD8$^{+}$ Tcells, NK cells, CD19$^{+}$B cells, naïve switch and unswitched B cells upon ld-IL2 treatment. Data are shown as shown as fold change from baseline

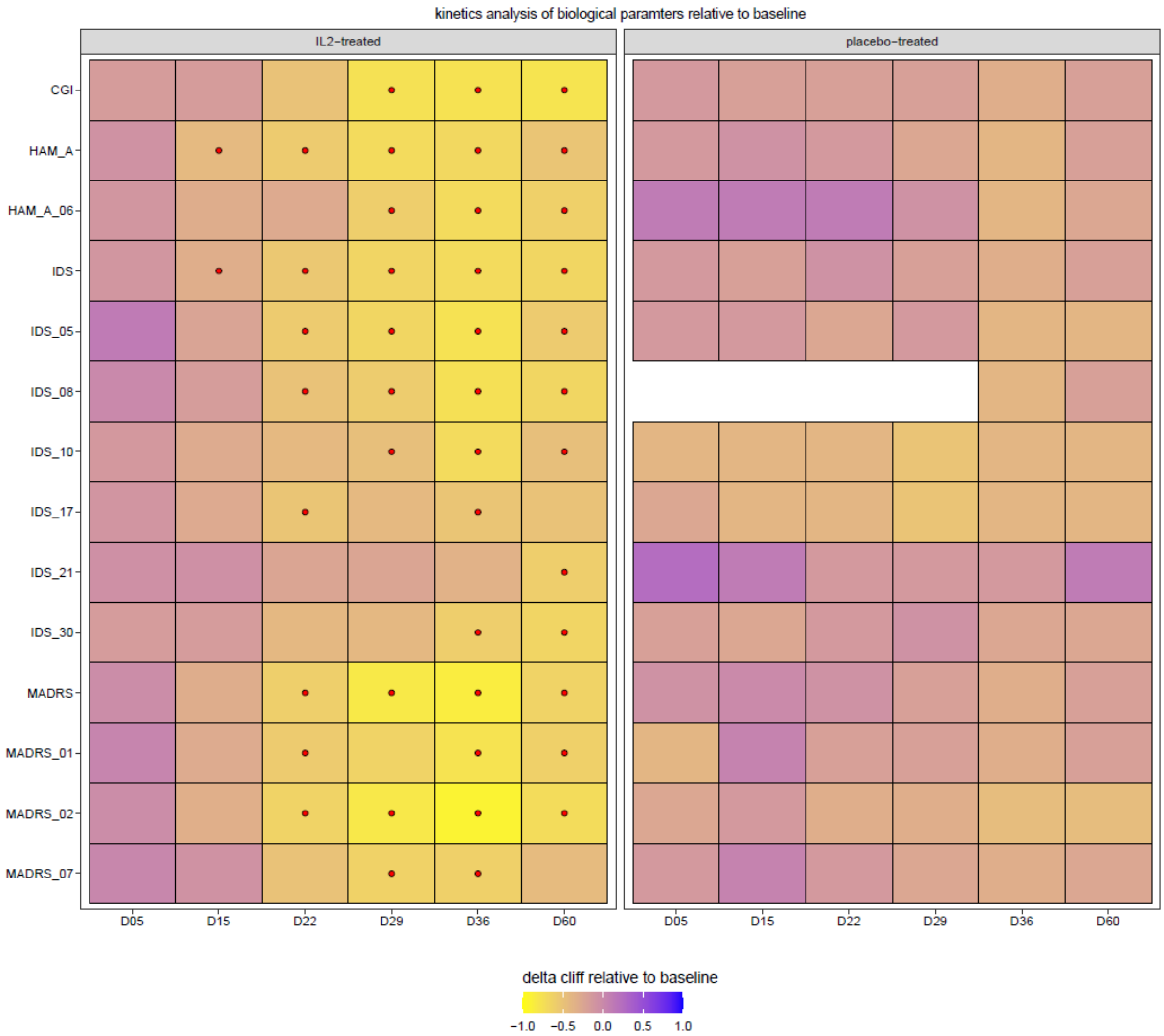


**Figure S4 – Heatmap representation of clinical scales and subscales modified along the time course of the study.**

The different clinical scales and subscales were compared to the baseline for IL-2 treated and placebo-treated patients. The magnitudes of changes are represented using the delta cliff by a color gradient scale ranging from -1 to 1. Statistically significant modifications are represented by a red dot.

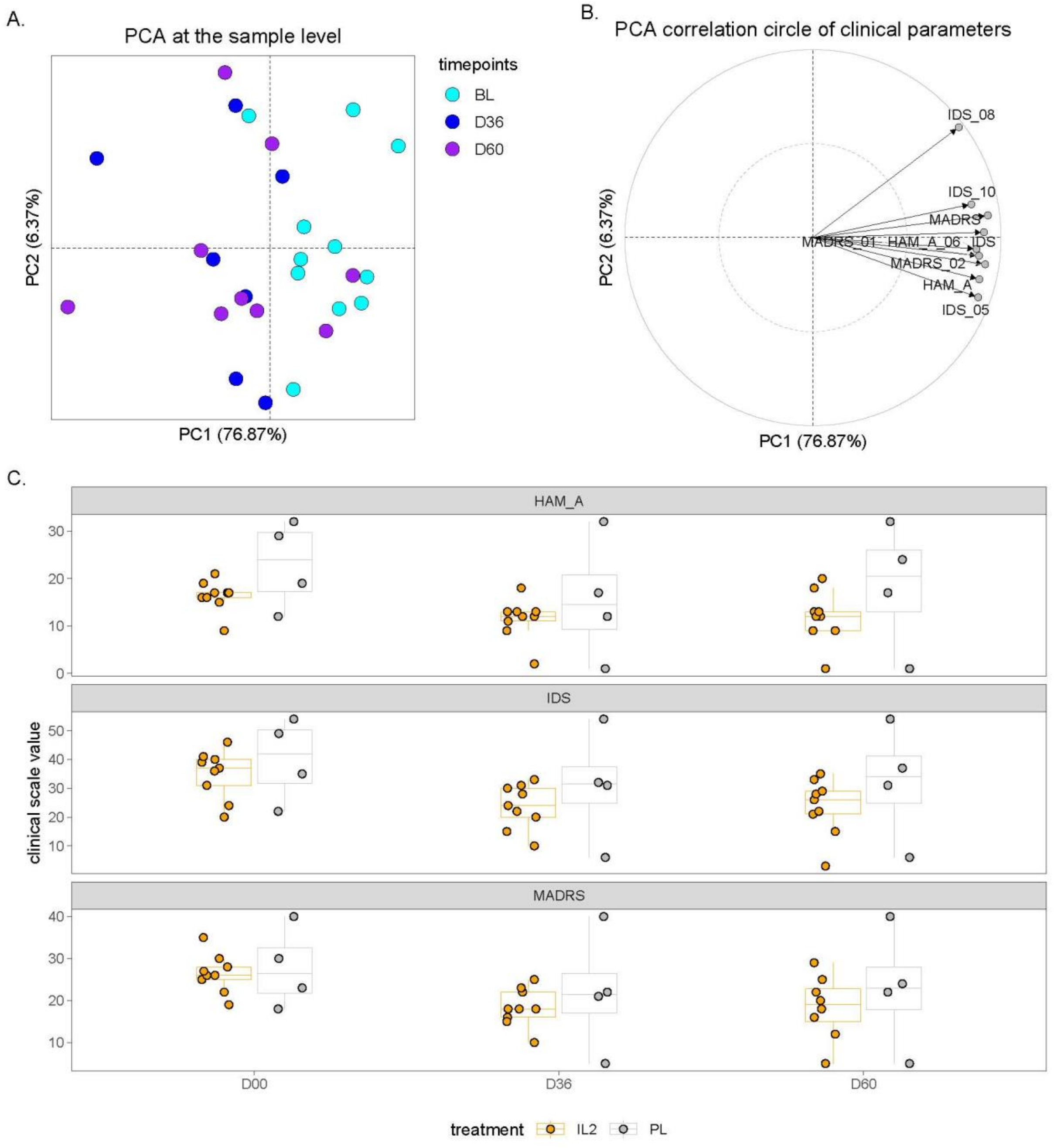


**Figure S5. Multivariate analysis of clinical changes during the treatment period and comparison with placebo-treated individuals**

**(A)** Principal component analysis of clinical scale at baseline, day 36, and day 60 post beginning of treatment for IL-2 treated patients. **(B)** PCA correlation circle showing clinical parameters associated with each axis of the PCA. (**C**) Jitter and boxplot representations showing the HAM-A, IDS, and MADRS changes between placebo and $IL2_{LD}$-treated individuals at baseline, D36, and D60 post-beginning of the treatment.

**Supplemental methods**

***Immunomonitoring***

Blood samples were obtained for specific immunological tests including assessment of Treg and lymphocyte subsets at day 1 (baseline), day 5 and for follow-up at day 60. The primary endpoint was the increase in the relative concentration of Treg cells, measured by flow cytometry as $CD3^{+}CD4^{+}CD25^{hi}CD127^{-/lo}FoxP3^{+}$ cells among the $CD4^{+}$ T cells[1,2].

, at the end of the induction period (day 5) compared to baseline.

***Flow cytometry***

Blood samples were collected in EDTA tubes according to the planned protocol: absolute numbers of lymphocyte subsets and Tregs were monitored at each patient visit: day-1 (baseline) before treatment and during IL-2 administration at day 5, day 15, day 29, day 57, day 113 and day169 of the follow-up. The last day follow up was 2 months post treatment at day 253.

Blood subsets ($CD3^{+}$, $CD4^{+}$, $CD8^{+}$ T lymphocytes, $CD19^{+}$ B lymphocytes and $CD3^{-}CD56^{+}$ NK cells) counts (cells/µl) were established from fresh blood samples using CYTO-STAT tetraCHROME kits with Flowcount fluorescents beads and tetra CXP software with a FC500 cytometer (Beckman Coulter) according to manufacturer's instructions.

Foxp3 labeling was performed with custom designed dry tubes (duraclone from Beckman Coulter) containing CD3-FITC, CD25-PE, CD127-PE-Cy7, Foxp3-AF647, CD8-KRO, CD8-PB antibodies as described already described[1]. Total blood (60 µL) sampled with anticoagulant was mixed with 6 µL of Perfix-NC R1 buffer, vortexed immediately for 2-3 seconds and incubated for 15 min at room temperature in the dark. Perfix-NC R2 buffer (360 µL) was added and, after homogenization, a 360 µL aliquot was transferred to a Duraclone tube. After vortexing for 10 seconds, tubes were incubated for 60 min at room temperature in the dark. PBS 1X (3 mL) was added to the tubes, incubated for 5 min at room temperature in the dark before centrifugation for 6 min at 250g. The supernatant was removed to leave the pellet dried and the cells were resuspended in 3 mL of 1X Perfix-NC R3 buffer prior to another 6-min centrifugation at 250g. The pellet was dried and resuspended in 300 µL of 1X R3 buffer. Tubes were protected from light and stored at 4°C until the acquisition on a cytometer within the next 24 h. Acquisition was performed on a Navios cytometer (Beckman Coulter) maintained daily according to the manufacturer's recommendations with Flow Check Pro and Flow Set Pro fluorospheres. Acquisitions were performed, respectively, using Navios software, and all analyses were done with Kaluza 1.3 software (Beckman Coulter)[1].

Stat5 phosphorylation: 100 µl of EDTA-anticoagulated whole blood or 500 000 PBMCs were added to Duraclone part 1 tubes containing dried anti-CD4-Pacific Blue antibody (A82789), anti-CD45-Krome Orange (B36294), anti-CD127-PE-Cy7 (A64618), anti-CD45RA-FITC (A07786) and anti-CD14-ECD (IM2707U), all from Beckman Coulter, and different doses of dried ILT-101 (0, 0.01, 0.16, 0.8, 4, 20, 100 or 500 UI/ml). After 10 minutes of incubation in the dark at RT, cells were washed with 3 ml of PBS 1X and centrifuged at 300G for 5 minutes. After removal of supernatants, 50 µl of R1 (Perfix EXPOSE (B26976, Beckman Coulter) fixative reagent) were added in each tube followed by a 10 min incubation (dark, RT), then 1 ml of R2 (Perfix EXPOSE permeabilizing reagent) for whole blood or 0.5 ml of R2 and 0.5 of FCS for PBMCs was added prior to a 10 min incubation (dark, RT). After a 5 min centrifugation at 300G, cells were washed in 3 ml of PBS 1X and centrifuged again (300G, 5 min). Cells were resuspended in 200 µl of R3 (Perfix EXPOSE staining buffer) and transferred into Duraclone part 2 tubes containing anti-pSTAT5-PE (B23139) and anti-FoxP3-AF647 (B30650), both from Beckman Coulter. After an incubation of 45 min (RT, in the dark), 3 ml of PBS were added and tubes were incubated for 5 min (RT, in the dark). After centrifugation (300G, 5 min), cells were resuspended in 3 ml of R4 buffer, and after a final centrifugation (300G, 5min), resuspended in 250 µl of R4 prior to acquisition. Data were collected on a Navios flow cytometer (Beckman Coulter) and data were analyzed using Kaluza Analysis Software (Beckman)[2]. Deep Immunophenotyping of Treg was performed as already described by Pitoiset et al[3].

***Cytokines***

The circulating serum levels of a panel of pro- and anti-inflammatory molecules reflecting the main inflammatory pathways were determined for the whole study sample. Serum concentrations of cytokines namely GM-CSF, IL-1α, IL-5, IL-7, IL-12/23p40, IL-15, IL-16, IL-17A, TNF-β, VEGF, IFN-γ, IL-1β, IL-2, IL-4, IL-6, IL-8, IL-10, IL-12p70, IL-13, TNF-α, AA, CRP, VCAM-1, ICAM-1 were measured using a MesoScale Discovery (MSD) human V-Plex electro-chemiluminescence assay (MSD. Rockville. Maryland. USA) according to manufacturer's recommendations. Inflammatory markers with more than 15% of missing data and those with more than 50% of concentrations outside the limit of detection (LLOD) were excluded from the analysis.
